# Reproducibility and sensitivity of 36 methods to quantify the SARS-CoV-2 genetic signal in raw wastewater: findings from an interlaboratory methods evaluation in the U.S.

**DOI:** 10.1101/2020.11.02.20221622

**Authors:** Brian M. Pecson, Emily Darby, Charles N. Haas, Yamrot Amha, Mitchel Bartolo, Richard Danielson, Yeggie Dearborn, George Di Giovanni, Christobel Ferguson, Stephanie Fevig, Erica Gaddis, Don Gray, George Lukasik, Bonnie Mull, Liana Olivas, Adam Olivieri, Yan Qu, SARS-CoV-2 Interlaboratory Consortium

**Author notes:** These authors contributed equally (Brian M. Pecson, Emily Darby).

## Abstract

In response to COVID-19, the international water community rapidly developed methods to quantify the SARS-CoV-2 genetic signal in untreated wastewater. Wastewater surveillance using such methods has the potential to complement clinical testing in assessing community health. This interlaboratory assessment evaluated the reproducibility and sensitivity of 36 standard operating procedures (SOPs), divided into eight method groups based on sample concentration approach and whether solids were removed. Two raw wastewater samples were collected in August 2020, amended with a matrix spike (betacoronavirus OC43), and distributed to 32 laboratories across the U.S. Replicate samples analyzed in accordance with the project’s quality assurance plan showed high reproducibility across the 36 SOPs: 80% of the recovery-corrected results fell within a band of +/- 1.15-log10 genome copies/L with higher reproducibility observed within a single SOP (standard deviation of 0.13-log10). The inclusion of a solids removal step and the selection of a concentration method did not show a clear, systematic impact on the recovery-corrected results. Other methodological variations (e.g., pasteurization, primer set selection, and use of RT-qPCR or RT-dPCR platforms) generally resulted in small differences compared to other sources of variability. These findings suggest that a variety of methods are capable of producing reproducible results, though the same SOP or laboratory should be selected to track SARS-CoV-2 trends at a given facility. The methods showed a 7-log10 range of recovery efficiency and limit of detection highlighting the importance of recovery correction and the need to consider method sensitivity when selecting methods for wastewater surveillance.

## 2 Introduction

The international water community responded rapidly to the onset of the COVID-19 pandemic by developing methods to measure SARS-CoV-2 genome concentrations in wastewater^1-3^. This effort was prompted by the identification of fecal shedding of SARS-CoV-2 in infected individuals^4-6^. As a result, wastewater surveillance has the potential to complement clinical testing by providing a broad observational assessment of the community’s health^3, 7^. Such knowledge could help guide public health agencies to identify and respond to outbreaks. Unlike clinical data—which may be biased toward the evaluation of symptomatic individuals—wastewater contains regular inputs from the entire population representing all stages of infection from symptomatic to pre-symptomatic to asymptomatic individuals. Furthermore, recent studies have shown that wastewater surveillance can provide an early warning of community infection, with wastewater concentrations spiking several days before identification through clinical testing^7-11^.

In April, 2020, The Water Research Foundation (WRF) hosted an international summit to evaluate the use of wastewater surveillance as an indicator of the distribution of COVID-19 in communities^12^. The participants identified two priority applications for the use of wastewater surveillance data: 1) tracking trends in occurrence and 2) assessing the degree of community prevalence. One of the prerequisites for these applications, however, is the identification of reliable, reproducible, and sensitive methods^10, 12, 13^. To help address this issue, this study performed an interlaboratory evaluation of 36 different methods used to assess the genetic signal of SARS-CoV-2 in untreated wastewater. The nationwide study included 32 U.S. laboratories from 19 different states each processing split samples of two different raw wastewaters emanating from populations known to have high levels of infection. The project sought to identify if and how the SARS-CoV-2 findings were impacted by multiple methodological differences such as sample concentration method, pasteurization pre-treatment, primer/probe selection, and solids removal steps. The effort did not intend to standardize a single method, but evaluate whether the existing methods provide sufficient reliability and reproducibility to track trends in occurrence and assess the prevalence of community infection.

## 3 Methods

### 3.1 Participating labs

The 32 participating laboratories included 17 academic labs, 6 commercial labs, 4 non-municipal government labs, 3 municipalities, and 2 manufacturers of molecular tests (Table 1). Prior to the interlaboratory study, many of the labs were engaged in on-going monitoring efforts across the country. The participating labs agreed to follow the project’s quality assurance project plan (QAPP) described below and process ten independent samples over a one-week period. The project QAPP is described in detail in this section in addition to an overview of the 36 individual Standard Operating Procedures (SOPs) evaluated in the study.

**Table 1.**
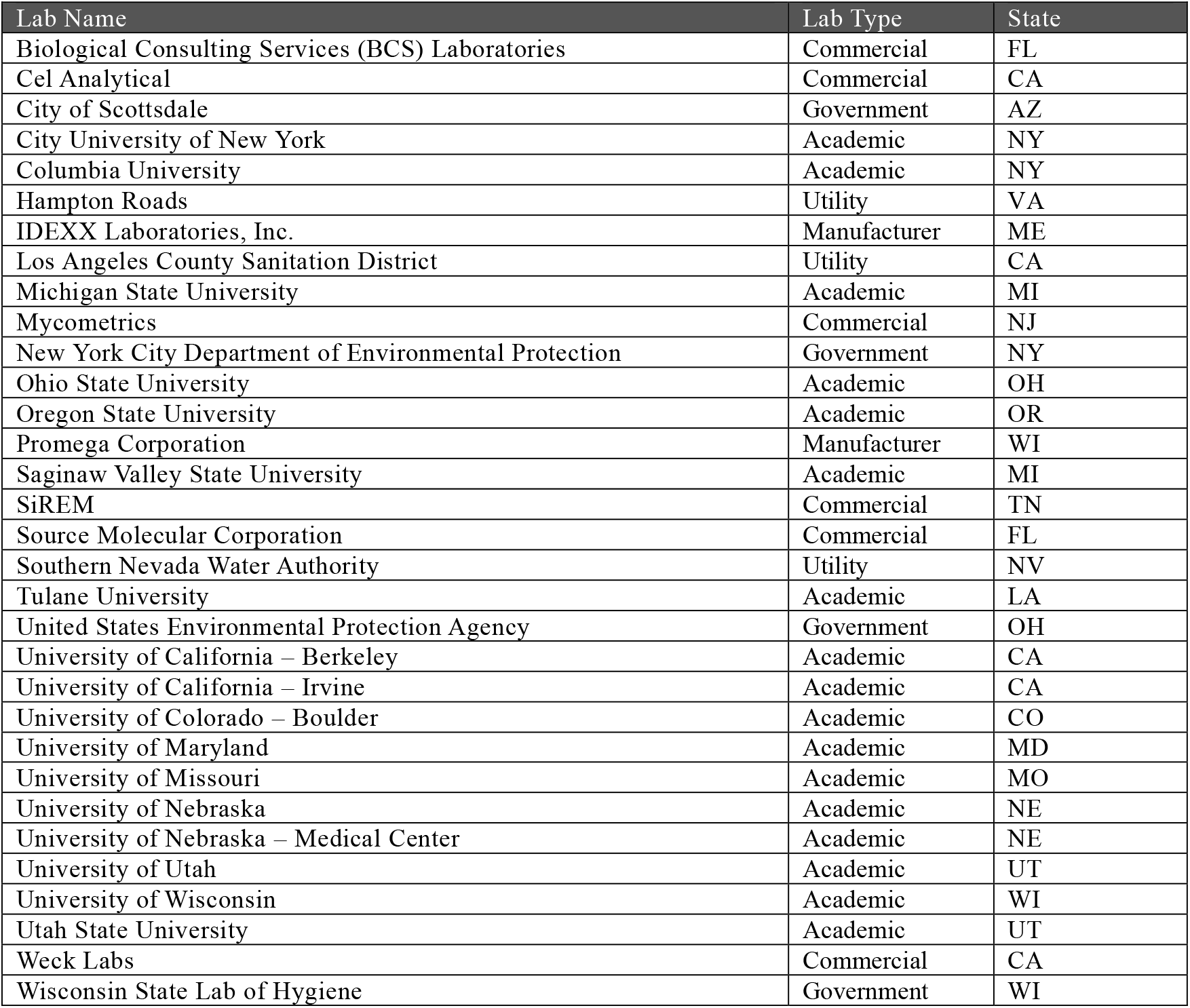
Participating Laboratories

### 3.2 Microorganisms

Human betacoronavirus OC43 was used as a matrix spike to assess the recovery efficiency of each method. To prepare the OC43 matrix spike, a concentrated stock of OC43 (Betacoronavirus 1 (ATCC® VR-1558™)) was grown in cell culture using HCT-8 cells (ATCC® CCL-244™), according to ATCC instructions. The concentration of OC43 genome copies (GC) in the stock was quantified by reverse transcription quantitative polymerase chain reaction (RT-qPCR) against a standard curve of Quantitative Genomic RNA from Betacoronavirus 1 (ATCC® VR-1558DQ™) to determine the GC per ml of the stock. Eight labs concurrently evaluated additional matrix spike organisms, including bovine coronavirus (BCoV), heat-inactivated SARS-CoV-2, bacteriophage MS2, bacteriophage Phi6, *in vitro* transcribed RNA, and an engineered RNA virus.

### 3.3 Sample collection, shipping, and handling

As detailed in the QAPP, raw wastewater samples were collected and distributed from two wastewater treatment plants (WWTPs) in Los Angeles County on two sampling days: (1) the Hyperion Water Reclamation Plant (operated by the City of Los Angeles Sanitation and Environment) on August 17, 2020 (Plant 1) and (2) the Joint Water Pollution Control Plant (operated by the Los Angeles County Sanitation Districts) on August 19, 2020 (Plant 2). These plants are two of the largest wastewater treatment plants on the west coast of the United States (Table 2). The sample collection location at both WWTPs was after grit removal prior to primary clarification. At both WWTPs, a single 40-gallon grab sample was collected at approximately 10:00 am. The bulk sample was distributed into 1-gal containers (one for each lab) while mixing the bulk sample continuously to promote homogeneity. To confirm the homogeneity of the samples, 1-L aliquots were collected after the 1^st^,17^th^,and 34^th^ samples and the total suspended solids, temperature, and pH were measured as surrogates for sample homogeneity (Table 2). The 1-gallon samples were chilled on dry ice to a temperature of approximately 4°C and then blind- spiked with betacoronavirus OC43 to a final concentration of 2.8 x 10^8^ GC/L. The samples were shipped to each laboratory with enough ice packs to maintain a temperature below 10°C. The participating labs were instructed to begin processing the sample between 8:00 AM and 12:00 PM Pacific time on the day after sample collection (i.e., 24 ± 2 hr after sample collection).

**Table 2.** WWTP Flows and Water Quality

| Parameter | Plant 1 | Plant 2 |
| --- | --- | --- |
| Annual Average Flow (MGD) | 275 | 260 |
| Total Suspended Solids (mg/L)<sup>1</sup> | 420 ($\pm 60$) | 520 ($\pm 40$) |
| pH<sup>1</sup> | 7.5 ($\pm 0.2$) | 6.9 ($\pm 0.1$) |
| Temperature (°C)<sup>1</sup> | 30 ($\pm 1$) | 38 ($\pm 1$) |

<sup>1</sup>Averages (plus/minus standard deviation) are based on the sample aliquots collected on the sampling day

### 3.4 Sample analysis

The participating labs each processed a total of 10 sample replicates. Most of the labs achieved these 10 sample replicates by processing five sample replicates from Plant 1 and five from Plant Eight laboratories evaluated the impact of heat pasteurization (60°C for 60 min) and so they achieved their 10 sample replicates by processing five sample replicates without heat pasteurization and five with heat pasteurization, all from Plant 1.

Each of the participating labs followed their own SOP for sample pre-treatment, concentration, extraction, and molecular analysis. Four of the participating labs tested two different SOPs leading to a total of 36 SOPs evaluated across the 32 labs. The SOPs were organized into eight method groups based on the concentration step prior to RNA extraction and whether solids were removed prior to concentration. The key method steps and categorization of the 36 SOPs are shown in Table Briefly, the starting sample volume ranged from 0.25 mL to 400 mL across the SOPs. The first step in sample processing was pre-treatment (e.g., heat pasteurization, solids removal, and/or chemical addition). Most labs did not pasteurize their samples before processing. SOPs involving heat pasteurization for all of the samples are marked with “H” and those involving heat pasteurization for half of the samples are marked with “(H)”. Approximately half of the SOPs involved the removal of solids (using either centrifugation, filtration, or both) prior to concentration. Method groups with SOPs involving solids removal are marked with an “S”. Many of the SOPs involved addition of chemicals to adjust the pH and/or the ionic composition of the matrix prior to concentration. After pre-treatment, the next major step in sample processing was concentration. The four main categories of concentration steps among these SOPs were 1) no concentration (i.e., direct extraction), 2) ultrafiltration, 3) filtration using an electronegative membrane (i.e., HA filter), and 4) PEG precipitation. The next step in sample processing was extraction. A variety of different extraction kits and in-house methods were used by the participating laboratories to extract the RNA from the sample. After extraction, the molecular analysis was conducted using either one-step or two-step RT-qPCR or reverse transcription digital PCR (RT-dPCR). All labs analyzed the native SARS-CoV-2 molecular signal using the N1 and N2 primer/probes sets and the OC43 matrix spike (Table 4). The concentration factors (CF) resulting from the different method steps of the SOPs, calculated using the equation below, ranged from 5 to 2100.

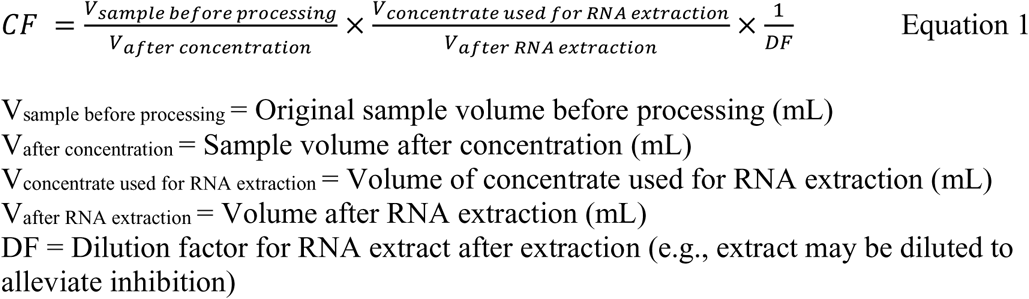

**Table 4.**
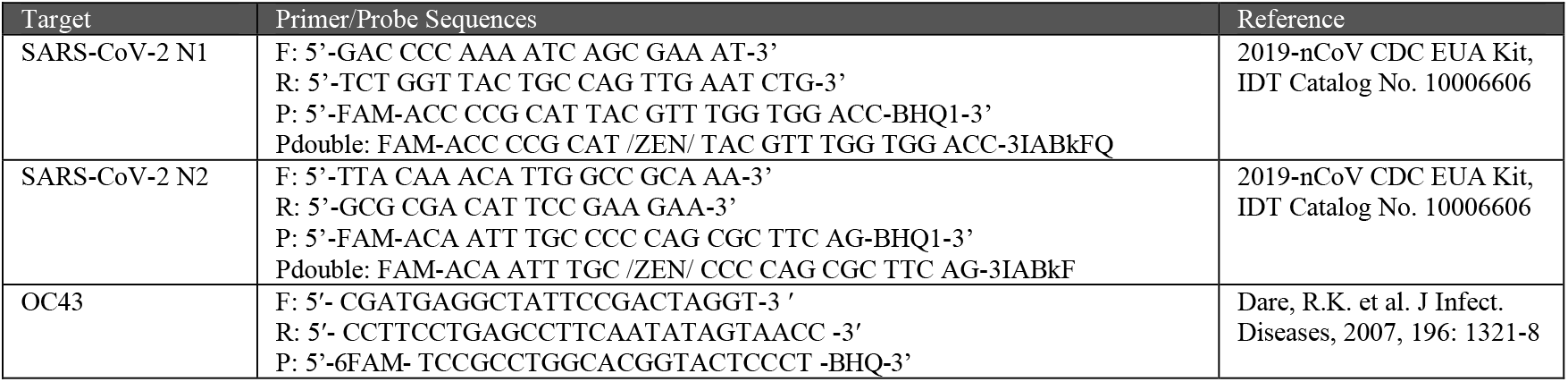
Primer and probe sequences for SARS-CoV-2 (N1 and N2 targets) and OC43

While each laboratory followed their own SOP, each lab was required to adhere to the project’s QAPP that described the quality control requirements^14^. The QAPP was constructed to ensure uniformity in sample collection, shipping and handling, quality control for the analytical methods, data management, and validation. Key elements of the QAPP included:

#### Blind matrix spikes

OC43 was spiked into each wastewater aliquot to achieve a final concentration of 2.8 x 10^8^ GC/L. The spike concentration was chosen to exceed typical background levels by orders of magnitude. Each lab was required to analyze OC43 concentrations in the same RNA extract used for SARS-CoV-2 quantification. Results from the OC43 blind matrix spikes were used to determine the recovery efficiency for each method.

#### RT-qPCR standard curves

Standard curves were required for each qPCR plate in which an environmental sample was quantified. The QAPP did not specify the use of a single type of standard due to cost and time constraints; however, it did specify that any plasmid-based standards be linearized prior to use.

#### Positive control

At least one positive control per target was run on each PCR plate to identify false negative results.

#### No template control (NTC)

The QAPP specified the inclusion of NTCs using PCR grade water processed by the same PCR steps as the sample. NTCs were required on every PCR plate to identify false positive results.

#### Laboratory method blank

At least one method blank (i.e., reagent water handled and processed by the same steps as the wastewater sample) was required for every round of samples.

#### Inhibition control

To assess the presence of inhibitory substances, the QAPP required that a molecular target not naturally present in the matrix be added to two qPCR wells in addition to the environmental RNA extract. The same target was added to two additional wells with PCR grade water. If the difference in RT-qPCR cycle numbers was greater than 1.0 between the two samples (i.e., the environmental extract and the PCR grade water), the labs were required to dilute and re-run the sample. For dPCR, the signal in the environmental sample was compared to the signal in the PCR grade water. If the ratio was less than 0.5, the labs were required to dilute and re-run the sample.

#### Molecular duplicates

For each replicate RNA extract, the molecular analysis was performed in duplicate.

#### Optional matrix spike

Nine of the laboratories evaluated a second matrix spike organism in addition to the QAPP-specified OC43 spike. The labs were required to spike the second surrogate to the raw wastewater samples at concentrations exceeding the background concentration. The sample was processed and analyzed for the surrogate in the same replicates used to analyze for the native SARS-CoV-2 and the spiked OC43.

### 3.5 Data analysis

The following quality control exclusion criteria were used to determine which data were included in the method analysis.

#### Limit of detection

Only results within the linear region of the standard curve were accepted as quantifiable results above the detection limit. An allowance of one CT (corresponding to an approximate two-fold decrease in concentration) was given when determining whether the results were within the range covered by the standard curve. Results that were lower than one CT of the lowest quantifiable standard were considered non-detects (NDs). Results that were self-reported by the laboratory as below the limit of detection or the limit of quantification were considered NDs. Two thirds of the SOPs had at least one molecular replicate that was marked as non-detect due to these criteria.

#### Non-detects

NDs were not included in the method analysis. If one of the molecular replicates for a sample replicate was non-detect and the other was above the detection limit (duplicates were performed for each sample replicate), only the result above the detection limit was used. If both molecular replicates were non-detect, the result for the sample replicate was non-detect. The number of sample replicates that were non-detect for both molecular replicates is presented in the results section.

#### Standard curves

If multiple replicates were performed for each standard, only the replicates with quantifiable results were used to develop the standard curve.

#### Sample hold time

If the sample was processed more than 24 hours outside of the specified 4-h processing window (8 a.m. to 12 p.m. Pacific Time on the day after sample collection), the results were not included in the method analysis. The results from one SOP (1S.1(H)) were excluded based on this criterion. Exceptions were made for two labs (SOPs 2.1 and 3.6) who immediately froze the samples upon receipt.

#### Contamination

If all of the NTCs or method blanks for N1, N2, or OC43 gave positive results, the results were excluded from the method analysis. This exclusion criterion applied to SOP 3.2 (but only for the N1 target).

#### Minimum recovery efficiency

If the recovery of the OC43 matrix spike was less than 0.01%, the SARS-CoV-2 results were excluded from the method analysis. The results from two SOPs (2S.1 and 3S.1) were excluded based on this criterion. Nevertheless, the limit of detection could still be calculated for these SOPs so their values were included in the method sensitivity analysis.

#### Cross-reactivity between BCoV and OC43

Several of the laboratories reported cross-reactivity between OC43 and their second matrix spike, BCoV. Further investigation showed that the OC43 primer/probes detected BCoV but not vice versa. This was confirmed *in vitro* through quantification of BCoV cDNA with the OC43 assay as well as *in silico* using NCBI BLAST. Because the BCoV was typically spiked at concentrations that were an order of magnitude lower than OC43 (SOPs 1S.2H, 2S.3, 3.4, 4S.3, and 4S.7) and because the current OC43 assay had lower sensitivity towards BCoV genome than the BCoV assay, the impact was deemed to be negligible (< 10%). In one case (SOP 3.5), the OC43 and BCoV concentrations were the same order of magnitude. No correction to the OC43 recovery was deemed necessary because the BCoV matrix spike led to an approximate two-fold increase in concentrations, whereas the recovery efficiencies ranged over several orders of magnitude.

#### Amplification plots

Five of the SOPs (1.1, 2S.2, 2S.3, 4S.2(H), 4S.7) had non-sigmoidal amplification plots for all of the sample replicates while the standards had the expected sigmoidal shape. The results from these SOPs were not excluded for this reason, but it should be noted that there may be greater error associated with these results since the results are more dependent on the fluorescence threshold selected for qPCR quantification. A non-sigmoidal amplification curve may be due to a level of matrix interference that was not detected by the inhibition control (all five SOPs passed their inhibition controls).

#### Number of replicates

While most laboratories processed five sample replicates per sample, four labs processed three replicates per sample (SOPs 1S.3(H), 2.1, 2.2, and 4S.8(H)), one lab processed one replicate per sample (SOP 4.4), and SOP 4S.5H processed eight replicates for the Plant 1 and ten sample replicates for Plant 2. All data were included in the analysis.

A summary of the results that were excluded from the analysis are presented in Table 5.

**Table 5.** Quality Control Rationale for Exclusion of SOPs

| SOPs Excluded from Method Analysis | Quality Control Rationale |
| --- | --- |
| 1S.1(H) | Processed more than 24 hr outside specified window |
| 3.2 (excluded N1 results only) | Positives in N1 NTC |
| 2S.1 (still included in method sensitivity analysis) | Low recovery (<0.01%) |
| 3S.1(still included in method sensitivity analysis) | Low recovery (<0.01%) |
| Two thirds of SOPs had at least one molecular replicate that were marked as non-detect due to the results falling outside of the range covered by the standard curve. NDs were not included in the analysis of SARS-CoV-2 results, but the SOPs were still included in method sensitivity analysis. |  |

After applying the exclusion criteria, the results of the sample replicates from each WWTP were analyzed separately. In the eight cases where an SOP was tested with and without pasteurization, the results were analyzed independently. When analyzing data by method group, only the five replicates without pasteurization were included in the statistical analysis of the method groups so as to not give extra weight to those SOPs.

### 3.6 Statistical analysis

The statistical analysis was performed in R using the log10-transform of the SARS-CoV-2 concentration, recovery efficiency, and limit of detection^15^. One-way ANOVA was used to compare the results of the eight method groups. A Tukey post-hoc test was used to perform multiple pair-wise comparisons. Comparisons with a p-value less than 0.05 were considered significant. Two-way ANOVA, with an interaction term, was used to evaluate the impact of different method steps, specifically, heat pasteurization, solids removal, primer/probe target, PCR platform, and matrix spike selection. Two-way ANOVA allows for the evaluation of two independent variables. The difference between the two levels of the second independent variable are calculated at each level of the first independent variable and averaged to determine if the difference is significant. For each of the method steps evaluated, the first independent variable was either the SOP or the concentration step and the second independent variable was the method step of interest: heat pasteurization, solids removal, primer/probe target, PCR platform, and matrix spike surrogate. The dependent variable was either the SARS-CoV-2 concentration or the matrix spike recovery. When the design was unbalanced, a type III sum of squares approach was used for two-way ANOVA.

## 4 Results

Over 2000 data points were produced from the interlaboratory analyses. This section addresses the reproducibility and sensitivity of the methods, both across all SOPs as well as within each of the eight major method groups. In addition, the impact of several other method steps—namely, pasteurization, primer/probe set, PCR platform, and matrix spike surrogate selection—were evaluated.

### 4.1 Reproducibility

The reproducibility of the methods was evaluated at three different levels: 1) across all method groups, 2) within each method group, and 3) within each SOP.

#### Across all Methods

To evaluate the variability of the SARS-CoV-2 concentrations measured by the different SOPs, the log-transformed N1 and N2 concentrations measured in the Plant 1 sample replicates (corrected for recovery based on the OC43 matrix spike) were plotted in a box plot (Figure 1). The data showing the uncorrected values can be found in the Electronic Supplementary Information (Figure ESI-1). The majority of the SOPs had sufficient sensitivity to obtain quantifiable results for most or all of the sample replicates performed for Plant 1 and Plant 2. Data that were below the detection limit or that did not pass the quality control criteria were not included in this evaluation. 36 SOPs at Plant 1 and 22 SOPs at Plant 2 passed the quality control criteria and had at least one sample replicate with detectable concentrations (where methods processed both with and without pasteurization were considered distinct SOPs). The variability, or reproducibility, of the different SOPs was quantified by calculating the range in which 80% of the data fell. The 10^th^ and 90^th^ percentile concentrations were 4.4-log and 6.7-log genome copies per liter (GC/L), respectively, for the combined N1 and N2 datasets (shown as dashed lines in Figure 1). In other words, 80% of the values from 36 different SOPs fell within a +/- 1.15-log band (2.3-log range). While a similar degree of reproducibility was observed at Plant 2, fewer SOPs were tested since those evaluating the impact of pasteurization only processed the Plant 1 sample and a greater percentage of the samples that were processed resulted in NDs (data not shown).

**Figure 1.**
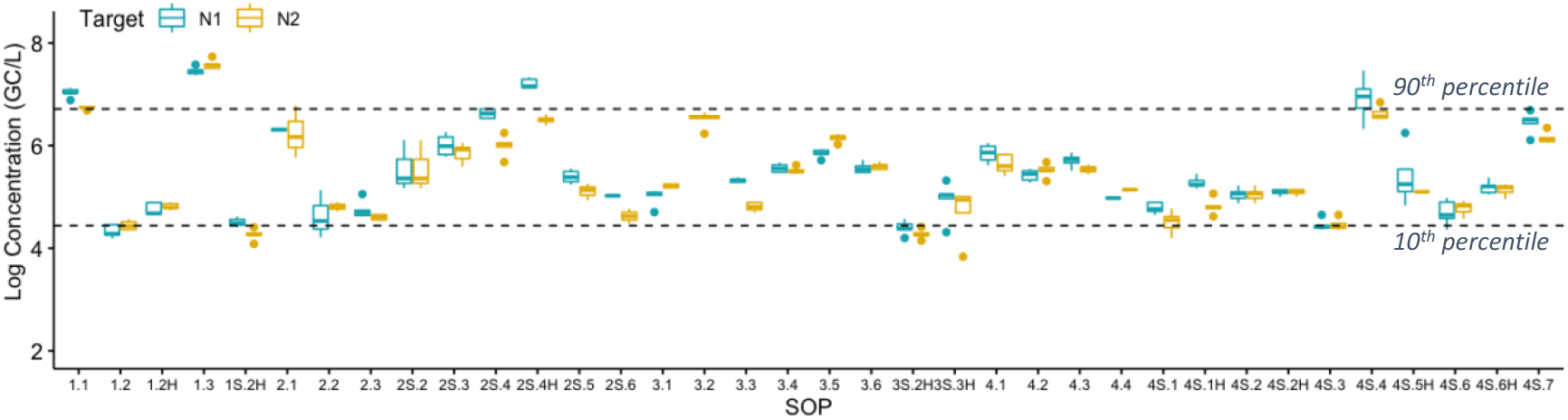
Recovery-corrected SARS-CoV-2 concentrations (N1 and N2 targets) at Plant 1 measured by each SOP. NDs and data excluded based on the quality control criteria are not plotted. The dashed lines show 10^th^ and 90^th^ percentiles across all N1 and N2 results.

In contrast, the recovery efficiency of the SOPs spanned seven orders of magnitude (Figure 2). Correcting for this source of methodological variability allowed the recovery-corrected concentrations to converge within a tighter minimum-maximum range than the uncorrected values (uncorrected data shown in Figure ESI-1), highlighting the importance of correcting for recovery in obtaining reproducible results across SOPs.

**Figure 2.**
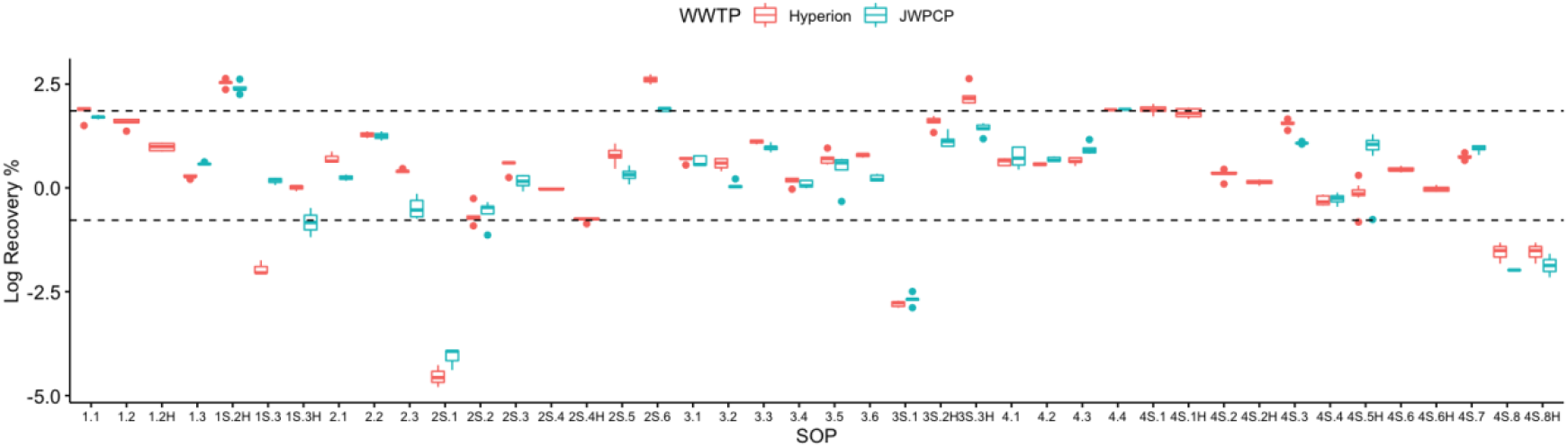
Log-transformed OC43 recovery efficiency at Plant 1 (Hyperion) and Plant 2 (JWPCP), measured by each SOP. The SARS-CoV-2 results from the SOPs highlighted are not represented in Figure 1 due to the fact that the results were all non-detect (gray), the recovery was below the quality control cut-off of 0.01% (blue), or both (orange).

#### Within a Method Group

The reproducibility of SOPs within each of the eight method groups was evaluated (Figure 3). The groups were based on the concentration step prior to RNA extraction—either (1) direct extraction or concentration by (2) ultrafiltration, (3) HA filtration, or (4) PEG precipitation—and whether solids were removed prior to concentration. The reproducibility within each method group was quantified by calculating the 10^th^ and 90^th^ percentile for the corrected SARS-CoV-2 concentrations from the replicates within each method group. Of the method groups with multiple SOPs, groups 3, 3S, and 4 had the greatest reproducibility with 10^th^-to-90^th^ percentile bands of 1-log or less. Method group 1 had the lowest reproducibility with a 10^th^-to-90^th^ percentile band of 3.2-logs. The factors leading to higher reproducibility within some method groups was not clear from the analysis. Potential factors include features inherent in the methods that lend themselves towards higher reproducibility or greater similarity of the SOPs within that method group. For example, three laboratories in method group 4 used a very similar SOP and had been in communication with each other prior to this study. The high reproducibility observed within group 4 suggests that aligning the details of an SOP between participants and greater interlaboratory communication may help to further improve the reproducibility of methods.

**Figure 3.**
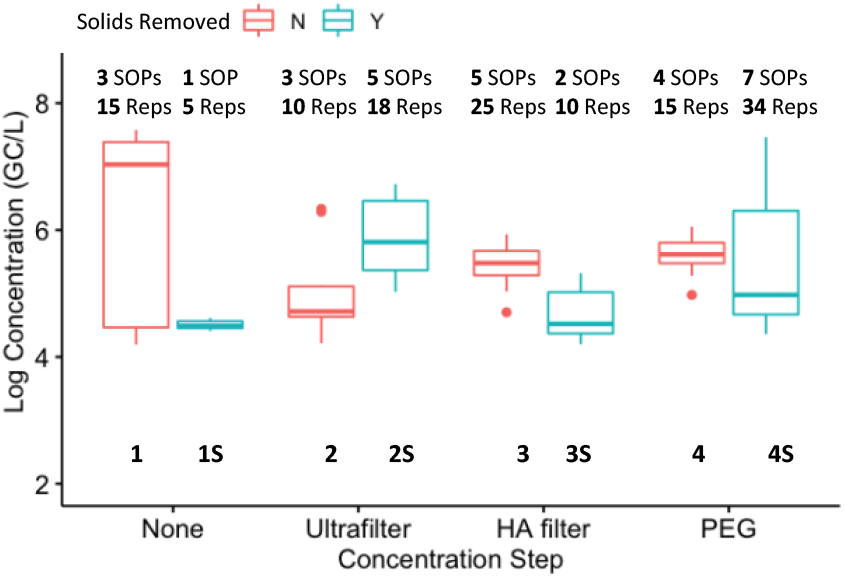
Comparison of the log-transformed SARS-CoV-2 (N1) concentrations at Plant 1 measured by each of the eight method groups (grouped by concentration step and solids removal). The number of SOPs and total sample replicates included in each method group are shown at the top of the box plot.

A box plot of the corrected SARS-CoV-2 N1 concentrations in eight method groups is shown in Figure 3. Given the variability of the pooled samples within the method groups, the recovery-corrected results from the different method groups were not systematically impacted by solids removal or concentration. Of the 28 pairwise combinations, only six had significant differences: 1S and 1 (p = 0.00047), 2 and 1 (p = 0.0028), 3 and 1 (p = 0.031), 4S and 1 (p = 0.0074), 2S and 1S (p = 0.013), and 3S and 2S (p = 0.0027). In other words, multiple methods led to similar results if the results were corrected for recovery. Similar trends were observed at Plant 2 (data not shown). Because only one or two SOPs were present in method groups 1S and 3S, the variability within those groups was not as well characterized as the other groups. Further studies with additional SOPs per group could be used to confirm the impact of solids removal and concentration steps.

#### Within each SOP

The reproducibility of each SOP was determined by calculating the standard deviation of the log-transformed results for the five replicates processed by the laboratory (Table 6). The precision of the SOPs was high based on a median standard deviation of 0.13 for both the N1 and N2 targets at Plant 1. The reproducibility with an SOP generally increased after correcting for recovery.

**Table 6.** Median and Range of Standard Deviations for Sample Replicates Processed by the Same SOP

| Target | Uncorrected | Recovery-Corrected |
| --- | --- | --- |
| N1 | **0.15** [0.04 – 0.38] | **0.13** [0.032 – 0.60] |
| N2 | **0.14** [0.01 – 0.53] | **0.13** [0.033 – 0.51] |

## 4.2 Sensitivity

The sensitivity of each SOP was evaluated by quantifying the theoretical limit of detection (LOD), which was, in turn, a function of three variables: the recovery efficiency, the concentration factor (CF), and the instrument detection limit of the PCR platform. The recovery efficiency for each SOP was calculated as the percentage of the OC43 matrix spike that was detected by the method (Figure 2). The concentration factor quantified the degree to which the SARS-CoV-2 concentrations increased as the raw wastewater was processed to produce the final RNA extract. Concentrations factors were SOP-dependent (Table 3). The instrument detection limit is the lowest concentration at which the PCR instrument can reliably distinguish a target signal from the background. Rigorous methods for quantifying instrument detection limits have been described previously^16^, but were not evaluated during this study. In lieu of this, a theoretical instrument detection limit of one GC per 5 µl PCR assay was assumed.

**Table 3.**
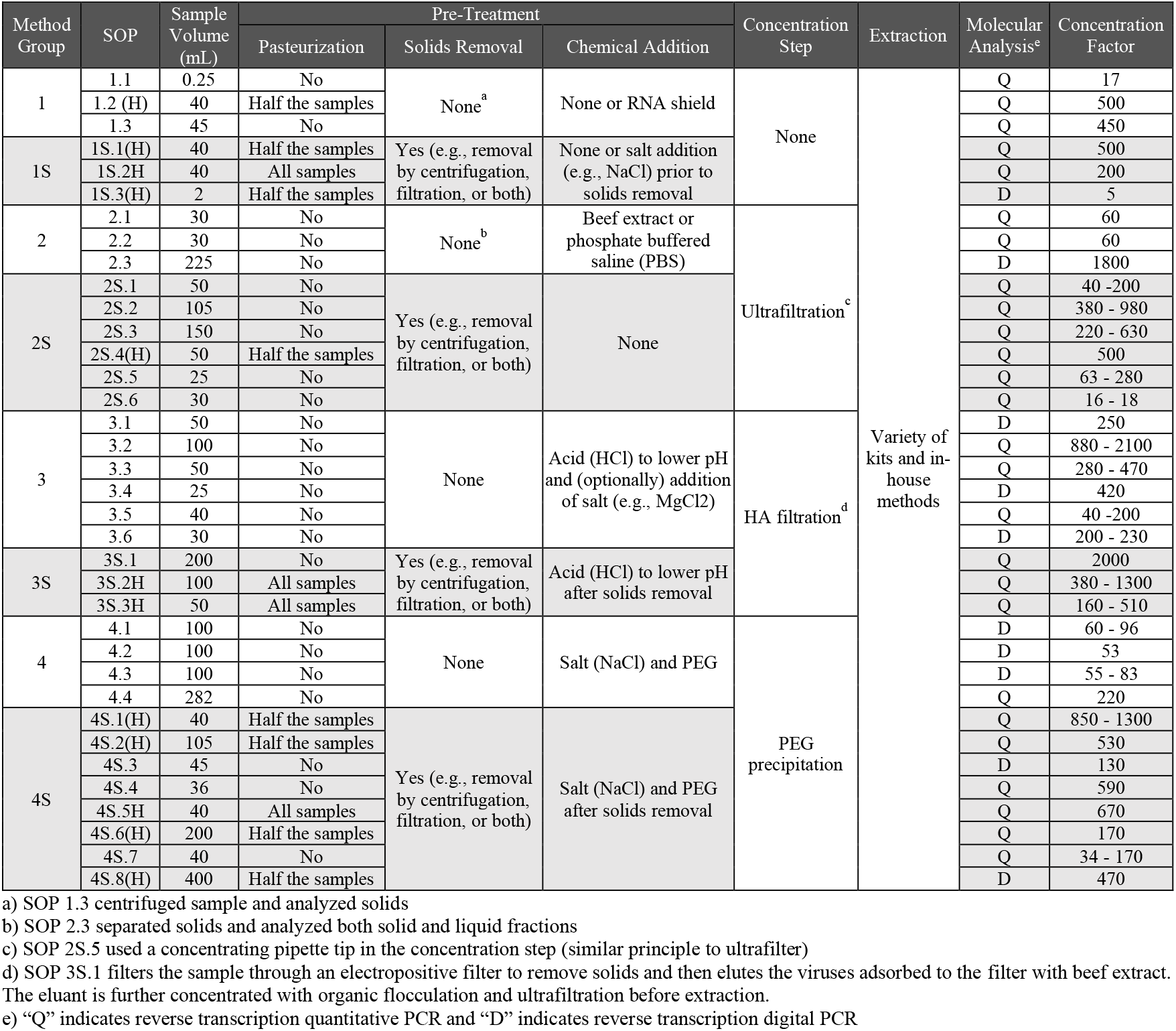
Key Method Steps and Categorization of the SOPs

These three factors were used to calculate the theoretical LOD for each SOP:

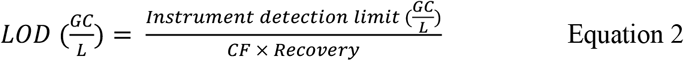

The theoretical LOD of the SOPs spanned more than six orders of magnitude (Figure 4). The high degree of variability in LODs was due largely to the recovery efficiencies, which also exhibited a similar range of magnitudes. The band defining the 10^th^ and 90^th^ percentiles spanned from a theoretical LOD of 3.0- to 6.1-log GC/L. To understand the sensitivity of the methods to detect lower concentrations than those present in the August 2020 wastewater samples, the log-difference between the measured SARS-CoV-2 concentrations and the theoretical LOD was determined for each SOP. The median difference across all methods was 0.8 logs, though some methods could detect concentrations 2-log lower or more.

**Figure 4.**
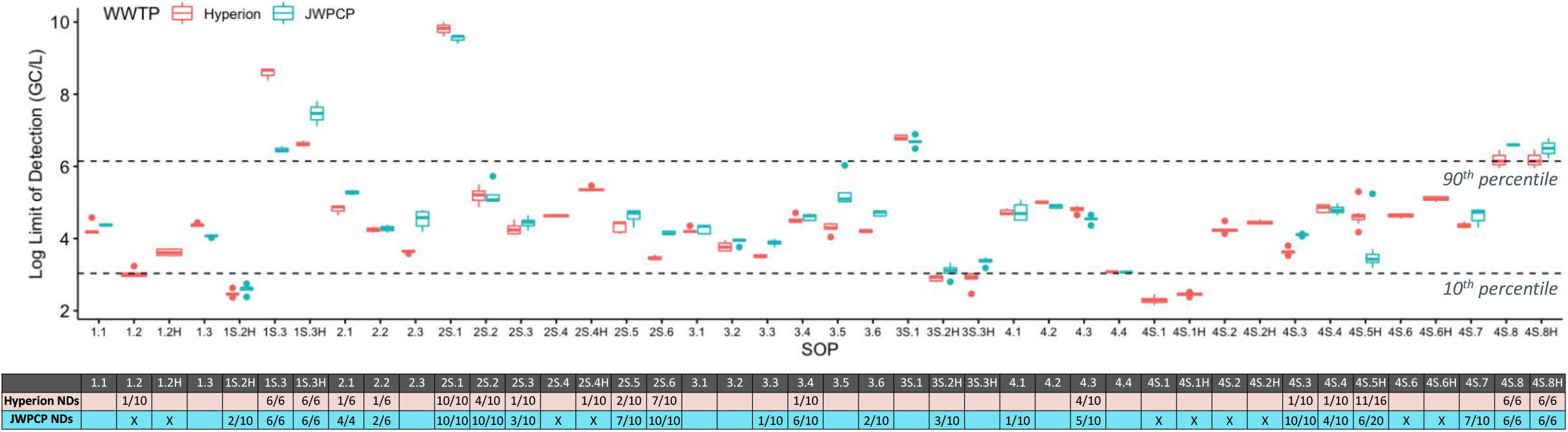
Log-transformed theoretical limits of detection for each SOP at Plant 1 (Hyperion) and Plant 2 (JWPCP). The dashed lines show 10^th^ and 90^th^ percentiles across both Plant 1 and Plant 2. The total number of non-detects (ND) (combined for SARS-CoV-2 N1 and N2 targets) out of total number of sample replicates processed by each SOP is shown in the table below the box plot (a blank cell indicates no NDs). An “X” indicates the sample was not processed by that SOP.

The variabilities in sensitivities can also be evaluated based on the frequency of sample replicates with NDs at each WWTP. As anticipated, SOPs with higher LODs (lower sensitivity) tended to have higher rates of NDs, and SOPs with lower LODs (higher sensitivity) tended to have fewer NDs (Figure 5). Recall, the theoretical LOD is based on the observed OC43 recovery—the actual SARS-CoV-2 recovery was not directly measured. Therefore, the fact that a strong relationship is observed between the LOD and the frequency of NDs suggests that OC43 is generally providing an accurate reflection of the *relative* SARS-CoV-2 recovery across different methods. It should be noted, however, that other factors affecting OC43 recovery at each lab (e.g., sample-to-sample differences, shipping effects, sample handling) may also contribute to the differences in the calculated LODs.

**Figure 5.**
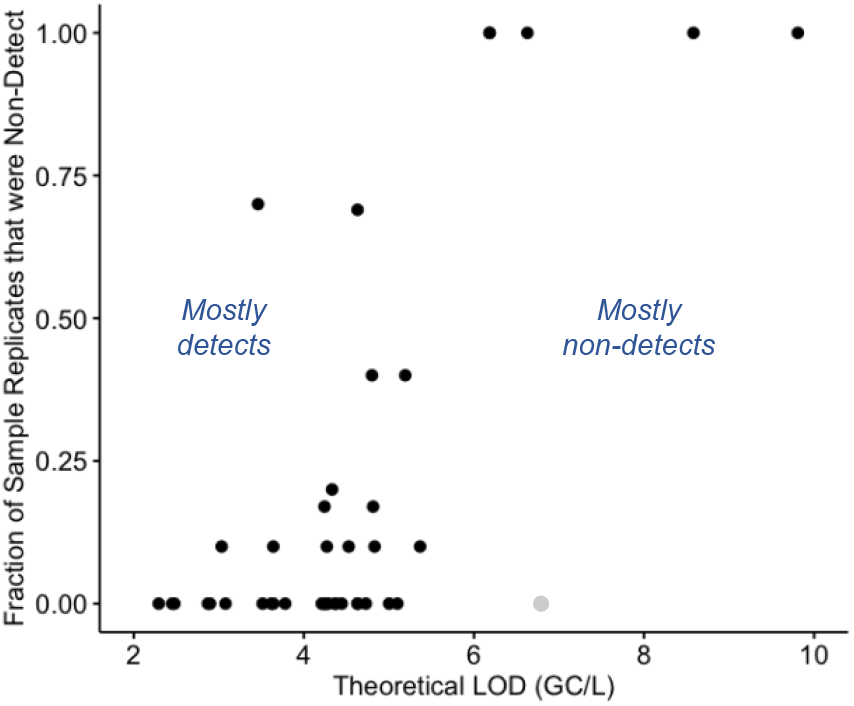
Fraction of sample replicates that were non-detect at Plant 1 as a function of the theoretical LOD. The outlier shown in gray (SOP 3S.1) processed the sample using a different PCR platform to enumerate OC43 and SARS-CoV-2.

To assess whether sensitivity was linked to methodological differences, the LODs for both WWTPs were compared by method group (Figure 6). The LODs between method groups was generally indistinguishable, partially due to the high variability of LODs within the method groups with solids removal. In each of these solids removal groups, the large LOD range was driven by a single SOP in the group with a high LOD, specifically, 1S.3(H), 2S.1, 3S.1, and 4S.8(H). These SOPs all had NDs and/or recovery below 0.01%. Only three of the 28 pairwise combinations were significantly different and all were associated with method group 2S: 2S and 1(p = 0.0011), 2S and 3 (p = 0.0062), and 2S and 4S (p = 0.011). The SOPs with highest sensitivity were not all associated with the same method group, meaning that multiple methods may be capable of achieving high sensitivities.

**Figure 6.**
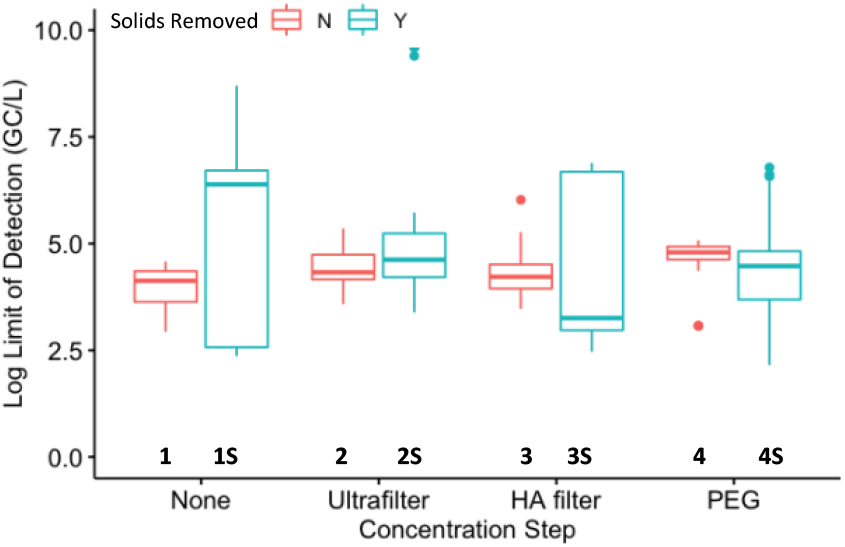
Comparison of the log-transformed theoretical limits of detection (combined for Plant 1 and Plant 2) for each of the eight method groups (grouped by concentration step and solids removal).

### 4.3 Impact of Other Method Steps

In addition to the main method steps differentiating the SOPs in this study (i.e., concentration step and solids removal), several other method steps were evaluated, namely heat pasteurization, primer set, PCR platform, and surrogate used as the matrix spike.

#### 4.3.1 Pasteurization

To evaluate whether heat pasteurization impacted the measured SARS-CoV-2 concentrations, five labs used their SOPs to process 10 replicates of the same wastewater: five without heat pasteurization and five with heat pasteurization conducted at 60°C for 60 min. Two-way ANOVA showed a statistically significant (p = 1.5 x 10^−13^) but small increase (0.41-log for N1 and 0.31-log for N2) in the corrected SARS-CoV-2 concentrations after pasteurization.

#### 4.3.2 Primer/probe set

To evaluate whether the selection of primer/probe set impacted the measured SARS-CoV-2 concentrations, all sample replicates were analyzed using both the N1 and N2 primer/probe sets. Two-way ANOVA showed a significant (p-value of 10^−8^ for Plant 1 and 0.00042 for Plant 2) but small difference between the results: N1 was 0.13-log greater than N2 at Plant 1 and 0.12-log greater at Plant 2.

#### 4.3.3 PCR Platform

To evaluate the impact of the PCR platform (quantitative PCR or digital PCR), the SOPs were grouped by platform within each method group (Figure 8). There was an unequal distribution of SOPs using quantitative and digital PCR across the different method groups. Of SOPs that passed the quality control and had detectable SARS-CoV-2 concentrations, 22 used quantitative PCR and eight used digital PCR; the eight SOPs that used digital PCR were distributed across only four of the method groups. The low sample numbers and unbalanced datasets made it difficult to perform a robust statistical comparison of the two platforms; based on the preliminary information, no clear patterns emerged between the two quantification platforms. Previous studies have indicated that dPCR may have advantages over qPCR in terms of increased sensitivity and resistance to inhibitory substances^17, 18^. Additional studies would be required to further evaluate the extent to which such differences exist for the SARS-CoV-2 methods.

**Figure 7.**
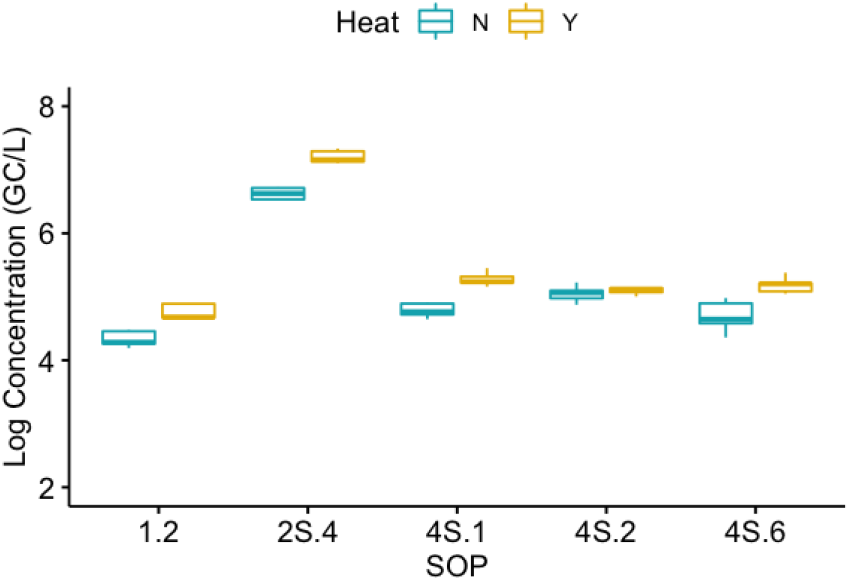
Impact of heat pasteurization on the log-transformed SARS-CoV-2 (N1 target) concentrations (corrected for recovery efficiency) at Plant 1. Five sample replicates for each SOP, with and without heat pasteurization, were performed.

**Figure 8.**
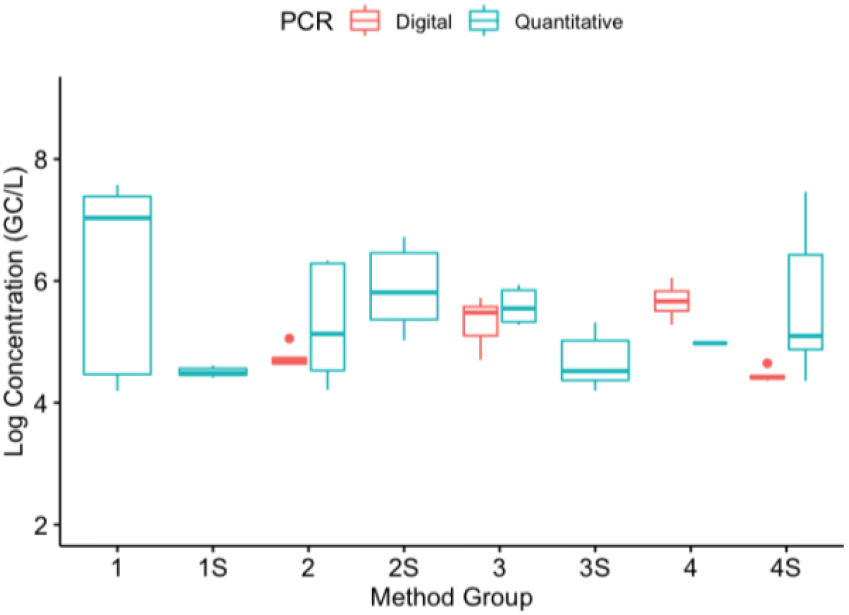
Impact of the PCR platform (digital or quantitative) on the log-transformed SARS-CoV-2 (N1 target) concentrations (corrected for recovery efficiency) at Plant 1. The data are from 22 SOPs (93 replicates) that used quantitative PCR and 8 SOPS (39 replicates) that used digital PCR.

#### 4.3.4 Matrix spike Selection used for Recovery Correction

The impact of matrix spike selection was evaluated by comparing the recovery of OC43 against a number of alternatives (Figure 9). All but one of the surrogates (i.e., in vitro transcribed RNA used in SOP 1.1) showed a statistically different recovery than OC43 (p <0.05), though the difference between OC43 and the other surrogates varied. For example, the difference between OC43 and the other betacoronaviruses—bovine coronavirus (BCoV) and heat-inactivated SARS-CoV-2—was relatively small compared to the other surrogates (average of 0.35-log and 0.47-log higher than OC43, respectively). One systematic difference was that OC43 was added upon sample collection before shipment to the labs whereas the second matrix spike was added upon receipt by the individual labs. A lower recovery for OC43 could be the result of decay that occurred in the sample during shipment that was not accounted for by the second surrogate. In comparison to the other betacoronaviruses, other surrogates had larger differences in recovery than OC43. For example, enveloped bacteriophage Phi6 had a recovery that was 3.9-log lower than the OC43 recovery. It is important to note that differences in surrogate recovery may be SOP-dependent, meaning that a surrogate may behave similarly to another in one SOP but differently in another. These findings suggest that multiple surrogates may be acceptable, but highlight the differences between some of the commonly used selections.

**Figure 9.**
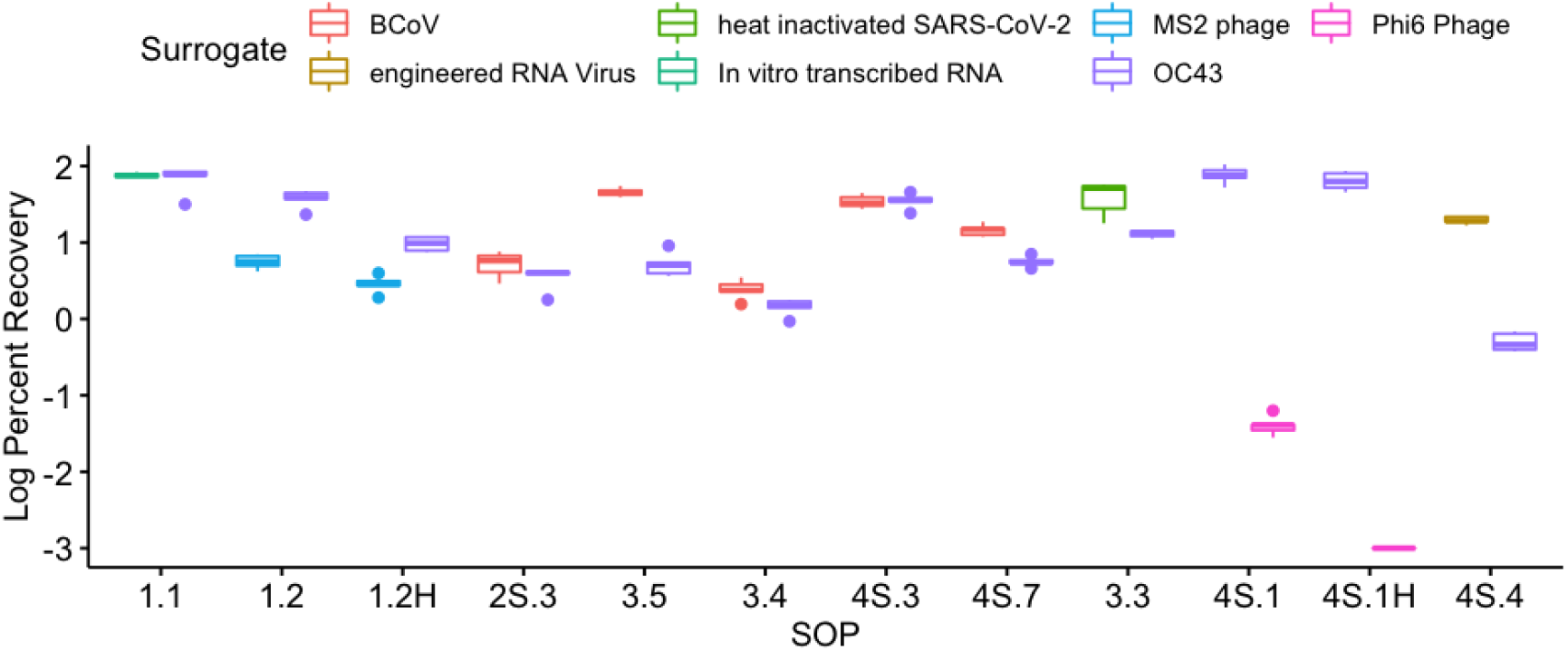
Impact of the surrogate used for the matrix spike on the log-transformed recovery efficiency at Plant 1. Five sample replicates for each SOP were processed and analyzed for both OC43 and the second matrix spike surrogate.

## 5 Discussion

This study demonstrated that a diverse set of 36 methods was able to quantify the SARS-CoV-2 genetic signal in raw wastewater with a high degree of reproducibility. 80% of the data from the eight different method groups fell within a band of approximately +/- 1-log GC/L. This finding bodes well for the nationwide interest in tracking SARS-CoV-2 in raw wastewater since a single standardized method may not be critical for obtaining comparable results between laboratories. Access to multiple, reliable methods may also increase the number of labs capable of participating in monitoring efforts and provide resilience against supply chain issues that have beset these efforts during the pandemic.

The findings also show, however, that methods-related hurdles remain before using the data for watershed-based epidemiology and modeling (e.g., estimating incidence and prevalence). This end use requires obtaining *accurate* information on the absolute concentration of SARS-CoV-2 genetic material in raw wastewater in addition to other information such as fecal shedding rates as noted below. Unfortunately, the accuracy of the methods—i.e., their ability to correctly quantify the true number of SARS-CoV-2 genome copies—could not be assessed because the actual concentrations in the raw wastewater samples were unknown. Despite the relatively tight band of results (80% within +/- 1 log), this 2-log range may be too wide for estimating community infection since 2 logs represents the difference between 1% and 100% of the population being infected. Additional data gaps must also be addressed for accurately modeling community infections including information on a) viral shedding rates in feces during different stages of infection^6, 19, 20^, b) how the genetic signal changes during travel through the wastewater collection system^21-23^, and c) sewershed modeling to estimate travel time and dilution. Multiple efforts should be pursued to address these knowledge gaps.

The findings are encouraging, however, for tracking *changes* or trends in virus concentrations. For this purpose, the absolute numbers quantified are not as important as identifying when and to what degree those numbers are increasing or decreasing^24^. The collection of SARS-CoV-2 wastewater concentrations could be used in conjunction with clinical data to provide complementary information on the extent of community infection and the effectiveness of public health interventions. The data could also be used to identify “hot spots” within a collection system where higher virus concentrations are measured ^7-9^. This knowledge could be used to trigger additional investigations of the populations within that sub-sewershed to identify and respond to communities experiencing higher infection rates. One benefit of this type of tracking is that the changes in wastewater concentrations may precede the clinical evidence of infection by multiple days, allowing for more responsive and focused public health interventions. A related use of this approach is confirmation of ongoing low community prevalence of SARS-CoV-2 in areas, such as small rural regions, for which testing rates are low. The use of wastewater surveillance as a sentinel for community infection has been described in Utah and at the University of Arizona^11^.

This study’s findings would suggest that the same method or laboratory be used to assess the SARS-CoV-2 concentrations over time at a given set of locations. For example, use Method A to assess trends within the sewersheds in Region X over time rather than switching between Methods A, B, and C over the monitoring period. Other regions (e.g., Region Y) could select different methods, but should then use the same method over the entire testing period to facilitate the tracking of trends. One exception to this may be cases in which multiple laboratories use a similar SOP and have demonstrated a high degree of reproducibility across labs, such as SOPs 4.1, 4.2, and 4.3. Given the high degree of intra-method reproducibility observed (standard deviation < 0.2 log GC/L), many methods have sufficient precision to sensitively detect *when* changes in virus concentrations are occurring. Collecting samples at multiple locations will also help identify where they are occurring.

### Factors promoting reproducibility

The high inter-method reproducibility was the result of three key factors: 1) the results were largely unaffected by methodological differences, 2) only data passing all QA/QC checks were included in the analysis, and 3) the QAPP normalized the findings to account for important sources of variability.

#### Minimal impact of methodological differences

The 36 methods were divided into eight groups based on two major methodological differences: the presence or absence of both a solids removal step and a sample concentration step. Based on this study’s findings, neither of these methodological branch points caused a clear, systematic impact on the enumeration of SARS-CoV-2 levels particularly after correcting for differences in recovery (see below). Additional work is recommended to further confirm these findings, though the preliminary data suggest that these differences are not important sources of variability.

Another positive finding was that the use of pasteurization prior to processing led to only modest impacts on virus enumeration when recovery correction was incorporated. This variability of approximately 0.3 to 0.4 logs may be acceptable, particularly if pasteurization pre-treatment is a requirement for lab safety. Multiple participants in the interlaboratory comparison noted that their institutions mandated pre-pasteurization (per CDC guidelines) to minimize the lab staffs’ exposure to the infectious agents in the raw wastewater (both SARS-CoV-2 and other pathogenic viruses and microorganisms). One concern was that pasteurization steps have been previously shown to impact both the infectivity and genetic signal of other viruses when heated at 72°C^25^. The QAPP prescribed lower temperature, longer duration conditions for pasteurization (60°C for 60 minutes) since it was hypothesized that higher temperature, shorter duration conditions may have a greater impact on virus fate^26-29^. Future studies could be used to confirm the range of acceptable pasteurization conditions for SARS-CoV-2 samples.

The two primer sets developed by the CDC for clinical diagnosis were used in this study. While the N1 primer set led to significantly higher concentrations than N2, these differences were considered to be minimal (approximately 0.1 log difference) compared to the other sources of variability. These findings suggest that future efforts may not need to evaluate both primer sets for tracking wastewater concentrations of SARS-CoV-2. Reducing the number of total PCR reactions per assay may be of particular interest for resource-constrained settings, though care should be taken to ensure that primer/probe sets account for mutational changes in the RNA sequence. The study also included methods using both qPCR and dPCR. Given the low number of dPCR methods evaluated, there was not sufficient statistical power to compare the results from the two platforms. Based on a preliminary analysis of the data, no clear pattern of differences emerged between the two quantification platforms suggesting both may be acceptable for future monitoring.

Moving forward, additional elements could be specified in the QAPP that may further improve the reproducibility across methods. For example, specifying the type of standards to be used and how the samples are shipped and stored prior to processing may further control variability. The high reproducibility between SOPs 4.1, 4.2, and 4.3 also suggests that greater consistency between SOPs and improved coordination between labs can further improve reproducibility.

#### Identification and selection of high-quality data

One of the key conclusions from this study is that any future monitoring efforts that entail the use of multiple methods should impose a minimum set of QA/QC requirements via a QAPP. The scope of the QAPP should cover the entirety of the process from sample collection, shipping, and handling, to acceptable analytical methods, to quality control requirements, data management, and validation. In this study, the QAPP ensured that all split samples were homogeneously distributed and processed within a narrow, specified window. This degree of detail was deemed critical to assess method reproducibility since some preliminary data suggested that the virus integrity may decay relatively rapidly with time and temperature^11^. Through the QA/QC requirements specified—including the use of non-template controls, extraction controls, matrix spikes, and qPCR standards—a handful of data were flagged and eliminated from the analysis (Table 5). By specifying these QA/QC requirements, data that failed these checks were identified and justifiably eliminated from the dataset, allowing the team to focus on methodological sources of variability.

#### Normalizing across methods

One benefit of a large interlaboratory method comparison is that it provides an opportunity to compare methods in a setting where many variables are held constant. One unexpected finding was the wide range of recovery efficiencies represented by the different methods. More than seven orders of magnitude separated the methods at the extremes indicating a more than 10 million-fold difference in their ability to recover the OC43 betacoronavirus from the wastewater matrix. Because of this huge range, correcting based on the matrix spike recovery was deemed critical since *not* correcting for this factor could lead to equivalent magnitudes of variability. This recommendation is in line with recent work by Li et al. (2019).

One challenge with correcting for recovery is that it assumes that the matrix spike behaves similarly to the target virus. Additional studies are needed to assess how well OC43 mimics SARS-CoV-2 behavior in wastewater matrices, meaning that correcting based on OC43 (or any other viral surrogate) may also introduce some degree of variability in the results. For example, differences between SARS-CoV-2 and the matrix spike organism in terms of solids association, thermal sensitivity, extraction efficiency and surface properties may lead to variability when correcting for recovery after solids removal steps, pasteurization, and concentration methods, respectively. Nevertheless, the differences between SARS-CoV-2 and OC43 are likely to have a smaller net impact on the results than differences in recovery efficiency. The similarity in recovery efficiencies of the three betacoronaviruses tested in this study (OC43, BCoV, and heat-inactivated SARS-CoV-2) provides some assurance that OC43 may behave in a similar fashion to SARS-CoV-2. In a post-study poll of the laboratory participants, 87% supported the practice of reporting and correcting for recovery efficiency. Additional work to confirm the selection of matrix spike organisms is recommended.

### Evolving the Methods

Demonstrating the high degree of reproducibility between methods is an important step because it confirms that multiple methods can be used to obtain similar results in these complex matrices. This does not mean, however, that all of the methods are equally suited for all future efforts. One of the most promising end uses for these methods is to track SARS-CoV- 2 concentrations in wastewater as a bellwether for community health. Ideally, methods employed for such uses would have both high *precision* to identify upward or downward trends in the data as well as high *sensitivity* to quantify concentrations in both epidemic (high community infection) and endemic (low community infection) settings. To understand how the sensitivity of these methods translates to potential application of this tool in endemic settings, the prevalence of COVID-19 in Los Angeles County at the time of sampling was estimated. Assuming infected individuals shed SARS-CoV-2 in in their feces for at least 27 days^6^, then 61,000 people with confirmed infections were shedding SARS-CoV-2 in the wastewater samples collected during the study^31^. In a population of ten million people, this corresponds to 1 in 160 people. At this level of community infection, nearly all of the methods were able to achieve quantifiable results of virus concentrations. The degree to which the concentration in the wastewater (and consequently the percent of the population infected) could decrease while still obtaining quantifiable numbers will vary across the methods.

The methods showed a sizable range of theoretical limits of detection with most falling in the 10^3^ to 10^6^ GC/L range (in comparison, the measured SARS-CoV-2 were generally in the range of 10^4^ to 10^6^ GC/L). Methods with theoretical LODs as low as 10^2^ GC/L were also identified that would offer a 10- to 1000-fold improvement over those methods. Additional studies should identify the methods best suited for tracking trends, particularly those that offer high precision, reproducibility, and sensitivity. As the call for more expansive state- and nationwide monitoring programs increases, methods that offer higher throughput and lower processing time may also rise to the top.

The findings can also be used to identify methods that are best suited for areas with greater resource constraints, including those without the financial, technical, and material resources available in large U.S. cities. Through this lens, methods that have lower material costs, fewer and simpler steps, and require less specialized knowledge could offer important advantages. For example, the direct extraction methods forego the use of downstream concentration steps eliminating the need for filtration devices, centrifuges, and additional chemicals. Consequently, these methods may be cheaper, faster, and easier to run. Further research is needed to show if these methods can also provide sufficient precision, reproducibility, and sensitivity, to be the methods of choice for the diversity of locations across the country and globe.

## 6 Conclusions

- A nationwide interlaboratory comparison of methods for the quantification of SARS-CoV-2 genetic signal in wastewater showed a high degree of reproducibility. 80% of the results from eight method groups (36 different methods) fell within a band of approximately +/- 1-log GC/L.
- Recovery-corrected results did not show a systematic impact from solids removal or concentration method used. Additional methods steps that were evaluated (e.g., pasteurization, primer set selection, and PCR platform) generally resulted in small differences compared to other sources of variability.
- Factors leading to greater interlaboratory reproducibility include a) the relative insensitivity of the findings to methodological differences, b) the implementation of strict QA/QC requirements, c) the use of a quality assurance project plan to normalize the findings and account for important sources of variability, and d) implementing a shared SOP among different laboratories.
- The findings support the use of wastewater surveillance for tracking trends in the concentrations of SARS-CoV-2 within communities. They also highlight methodological challenges related to modeling incidence and prevalence.
- Additional metrics should be used to select the best methods for future efforts including method sensitivity, cost, equipment requirements, and simplicity.

## Data Availability

The data supporting the findings of this study are available within the article and its supplementary materials

## 8 Disclaimer

This manuscript has been reviewed by the U.S. EPA and approved for publication. Approval does not signify that the contents reflect the views of the Agency, nor does mention of trade names or commercial products constitute endorsement or recommendation for use.

## 9 Acknowledgements

The authors would like to thank The Water Research Foundation (project No. 5089) and the Bill & Melinda Gates Foundation for funding this research. We thank Mia Mattioli (Centers for Disease Control and Prevention) for her guidance on the project advisory committee, and Hunter Johnson and Mark Keller (interns at Trussell Technologies) for support with the project planning and sample collection. We thank the City of Los Angeles Sanitation and Environment and the Los Angeles County Sanitation District for supporting the sample collection.

## 10 Additional Information

### Group author details: SARS-CoV-2 Interlaboratory Consortium

Tiong Gim Aw: Environmental Health Sciences, School of Public Health and Tropical Medicine, Tulane University, New Orleans, LA; Nichole E. Brinkman: Office of Research and Development, U.S. Environmental Protection Agency, Cincinnati, OH; Kartik Chandran: Earth and Environmental Engineering, Columbia University, New York, NY; Francoise Chauvin: Bureau of Wastewater Treatment, New York City Department of Environmental Protection, Flushing, NY; John J. Dennehy: Biology, Queens College and The Graduate Center of The City University of New York, Queens, NY; Phil Dennis: SiREM Laboratory, Guelph, ON; Shuchen Feng: School of Freshwater Sciences, University of Wisconsin-Milwaukee, Milwaukee, WI; Matthew T. Flood: Fisheries and Wildlife, Michigan State University, East Lansing, MI; Raul Gonzalez: Hampton Roads Sanitation District, Virginia Beach, VA; Joe Hernandez: Microbiology, City of Scottsdale Water Campus, Scottsdale, AZ; Kayley H. Janssen: Wisconsin State Laboratory of Hygiene, University of Wisconsin-Madison, Madison, WI; Sunny Jiang: Civil and Environmental Engineering, University of California - Irvine, Irvine, CA; Marc C. Johnson: Molecular Microbiology and Immunology, University of Missouri, Columbia, MO; Devrim Kaya: Civil and Environmental Engineering, University of Maryland - College Park, College Park, MD; Huiling R. Lee: Mycometrics, LLC, Monmouth Jct, NJ; Jiyoung Lee: Division of Environmental Health Sciences, College of Public Health & Department of Food Science and Technology, Ohio State University, Columbus, OH; Xu Li: Civil and Environmental Engineering, University of Nebraska-Lincoln, Lincoln, NE; Cresten Mansfeldt: Civil. Environmental, and Architectural Engineering, University of Colorado – Boulder, Boulder, CO; Subhanjan Mondal: Promega Corporation, Fitchburg, WI; Kara L Nelson: Civil and Environmental Engineering, University of California – Berkeley, Berkeley, CA; Katerina Papp: Applied Research and Development Center, Southern Nevada Water Authority, Las Vegas, NV; Agustin E. Pierri: Weck Laboratories, Inc., Industry, CA; Catherine B. Pratt: College of Public Health, University of Nebraska Medical Center, Omaha, NE; Anda Quintero: Source Molecular Corporation, Miami Lakes, FL; Tyler Radniecki: School of Chemical, Biological and Environmental Engineering, Oregon State University, Corvallis, OR; Ryan A. Reinke: Microbiology, Los Angeles County Sanitation Districts, Whittier, CA; D. Keith Roper: Biological Engineering, Utah State University, Logan, UT; Tami L. Sivy: Chemistry, Saginaw Valley State University, University Center, MI; Brian M. Swalla: IDEXX Laboratories, Inc., Westbrook, ME; Jennifer Weidhaas: Civil and Environmental Engineering, University of Utah, Salt Lake City, UT.

### Statement of Author Contributions

Brian Pecson and Emily Darby developed the project plan, analyzed the data, and wrote the manuscript; Charles Haas participated in the statistical analysis, interpretation, and presentation of the findings; Yamrot Amha, Liana Olivas, and Yan Qu were responsible for the collection, preparation, and distribution of the wastewater samples; Mitchel Bartolo supported the study planning and data analysis; Richard Danielson and Yeggie Dearborn provided input on the project’s Quality Assurance Project Plan and processed the wastewater samples alongside the SARS-CoV-2 Interlaboratory Consortium; George Lukasik and Bonnie Mull prepared the matrix spike and other shared reagents, provided input on the project’s Quality Assurance Project Plan, and processed the wastewater samples alongside the SARS-CoV-2 Interlaboratory Consortium; George Di Giovanni, Erica Gaddis, Don Gray, and Adam Olivieri were members of the WRF Project Advisory Committee that advised on project planning and data interpretation; Christobel Ferguson and Stephanie Fevig provided coordination and organizational support between the research team, Project Advisory Committee, and laboratories; the SARS-CoV-2 Interlaboratory Consortium (group author list) processed the wastewater samples.

