## Supplementary Information for "Reproducibility and sensitivity of 36 methods to quantify the SARS-CoV-2 genetic signal in raw wastewater: findings from an interlaboratory methods evaluation in the U.S."

### Electronic Supplementary Information

The uncorrected SARS-CoV-2 concentrations, recovery efficiency, and frequency of non-detects at Plant 1 are presented in Figure ESI-1.

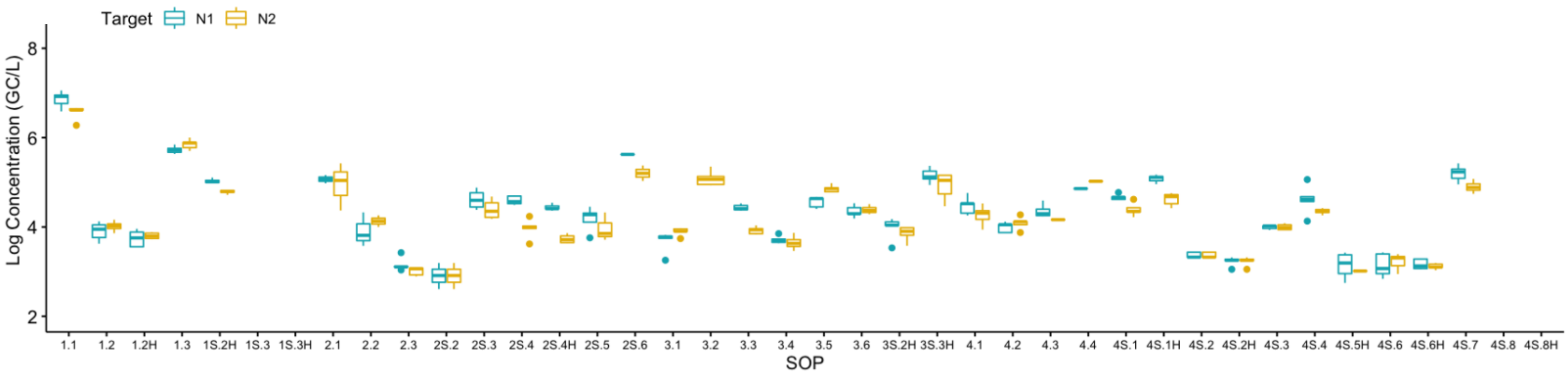


| | 1.1 | 1.2 | 1.2H | 1.3 | 1S.2H | 1S.3 | 1S.3H | 2.1 | 2.2 | 2.3 | 2S.2 | 2S.3 | 2S.4 | 2S.4H | 2S.5 | 2S.6 | 3.1 | 3.2 | 3.3 |
|---|---|---|---|---|---|---|---|---|---|---|---|---|---|---|---|---|---|---|---|
| N1 | | | | | | 3/3 | 3/3 | 1/3 | | | 2/5 | 1/5 | | | | 4/5 | | X | |
| N2 | | 1/5 | | | | 3/3 | 3/3 | | 1/3 | | 2/5 | | | 1/5 | 2/5 | 3/5 | | | |
| Average OC43 Recovery % | 71 | 38 | 10 | 1.9 | 340 | 0.01 | 1.0 | 5.3 | 19 | 2.6 | 0.25 | 3.6 | 0.94 | 0.17 | 6.8 | 410 | 4.8 | 3.9 | 13 |

| | 3.4 | 3.5 | 3.6 | 3S.2H | 3S.3H | 4.1 | 4.2 | 4.3 | 4.4 | 4S.1 | 4S.1H | 4S.2 | 4S.2H | 4S.3 | 4S.4 | 4S.5H | 4S.6 | 4S.6H | 4S.7 | 4S.8 | 4S.8H |
|---|---|---|---|---|---|---|---|---|---|---|---|---|---|---|---|---|---|---|---|---|---|
| N1 | | | | | | | | 1/5 | | | | | | | | 4/8 | | | | 3/3 | 3/3 |
| N2 | 1/5 | | | | | | | 3/5 | | | | | | 1/5 | 1/5 | 7/8 | | | | 3/3 | 3/3 |
| Average OC43 Recovery % | 1.5 | 5.4 | 6.2 | 40 | 190 | 4.3 | 3.7 | 4.5 | 76 | 78 | 65 | 2.2 | 1.4 | 36 | 0.51 | 0.85 | 2.8 | 0.96 | 5.7 | 0.03 | 0.03 |

**Figure ESI-1.** Uncorrected SARS-CoV-2 concentrations (N1 and N2) at Plant 1. The total number of non-detects (ND) (combined for SARS-CoV-2 N1 and N2 targets) out of total number of sample replicates processed by each SOP and the average OC43 recovery are shown in the table below the box plot (a blank cell indicates no non-detects).
